# The current quality and requirements of prehospital and emergency care in Indonesia and Malaysia: a cross-sectional multicenter survey

**DOI:** 10.1101/2023.08.20.23293872

**Authors:** Akio Tokita, Hanako Nunokawa, Keibun Liu, Yuta Iwamoto, Tomohiro Sonoo, Konan Hara, Mikio Nakajima, Kiyomitsu Fukaguchi, Takanori Takeda, Amirudin Sanip, Dafsah A. Juzar, Gurjeet Singh a/l Harvendhar Singh, Lukito Condro, Monalisa Tobing, Muhammad Abdus-Syakur bin Abu Hasan, Nik Hisamuddin NA Rahman, Orizanov Mahisa, Ramdinal Aviesena Zairinal, Mohd Khairulizwan bin Ramli, Mohd Afiq Mohd Nor, Tadahiro Goto, Mohd Idzwan bin Zakaria

**Affiliations:** School of Medicine, Faculty of Medicine, Gunma University, Gunma, Japan; School of Medicine, Tokyo Medical and Dental University, Tokyo, Japan; TXP Medical Co. Ltd., Tokyo, Japan; Department of Economics, University of Arizona, Tucson, AZ, USA; Emergency Life-Saving Technique Academy of Tokyo, Foundation for Ambulance Service Development, Tokyo, Japan; Department of Emergency Medicine, Shonan Kamakura General Hospital, Kanagawa, Japan; Department of Emergency Medicine, Universiti Kebangsaan Malaysia, Kuala Lumpur, Malaysia; Department of Cardiology and Vascular Medicine, Faculty of Medicine Universitas Indonesia - National Cardiovascular Center Harapan Kita, Jakarta, Indonesia; Hospital Selayang, Batu Caves, Malaysia; RSUD Kanjuruhan Kabupaten Malang, Kepanjen, Indonesia; Emergency Department, Hermina Depok Hospital, Depok, Indonesia; Kulliyyah of Medicine, International Islamic University Malaysia, Kuantan, Malaysia; Hospital Universiti Sains Malaysia, Health Campus, Kubang Kerian, Kota Bharu Kelantan, Malaysia; Muhammadiyah Hospital Lamongan, Lamongan, Indonesia; Head of Emergency Unit, Universitas Indonesia Hospital, Depok, Indonesia AND Department of Neurology, Faculty of Medicine, Universitas Indonesia, Jakarta, Indonesia; Hospital Tengku Ampun Afzan, Kuantan, Malaysia; Department of Emergency Medicine, University Malaya Medical Center, Kuala Lumpur, Malaysia; Academic Unit Trauma and Emergency, Faculty of Medicine, University Malaya, Kuala Lumpur, Malaysia

**Author notes:** Corresponding: Tadahiro Goto, MD, MPH, PhD TXP Medical Co. Ltd., Tokyo, Japan. Address: 7-3-1 Hongo, Bunkyo-ku, Tokyo, 113-0033. Akio Tokita and Hanako Nunokawa are equally contributed. Tadahiro Goto and Mohd Idzwan bin Zakaria are equally contributed.

**Keywords:** Ambulance, emergency room, Indonesia, Malaysia, quality improvement

## Abstract

**Background:** Rapid economic growth in Southeast Asian countries has led to a significant gap between supply and demand of emergency care, which may negatively affect the health outcomes of local populations. The purpose of this study was to identify current challenges faced by emergency care systems in Indonesia and Malaysia.

**Methods:** An online survey was conducted between August and November 2022. Survey participants consisted of emergency department (ED) doctors, nurses, and other medical staff in 11 hospitals in Indonesia and Malaysia. The survey collected information on respondents’ characteristics, factors associated with the quality of prehospital or ED care, missing clinical information, and factors associated with patients’ length of stay at the ED.

**Results:** A total of 83 and 109 respondents from Indonesia and Malaysia, respectively, answered the survey. The most important factor affecting prehospital care quality in both countries was “inadequate amount of clinical information from the ambulance.” The most important factor affecting ED care quality was “crowdedness in the emergency room during night shifts.” The clinical information most frequently missing was family history, followed by estimated time of arrival to the hospital or medication history. The primary factor affecting the length of ED stay was diagnostic studies and their turnaround time.

**Conclusions:** This study identified common challenges in the emergency care systems of Indonesia and Malaysia. Our findings highlight the importance of recognizing both common and country-specific challenges to improve the quality of emergency care in Southeast Asia.

## INTRODUCTION

Recent rapid economic growth in developing countries in Southeast Asia has resulted in a significant discrepancy between the supply and demand of emergency care, potentially leading to poorer health outcomes.[1] For example, mortality rates due to traffic accidents per 100,000 population are significantly higher in Indonesia (12.2) and Malaysia (23.6) compared to other developed countries in the Asia-Pacific, such as Singapore (2.8), Australia (5.6), and Japan (4.1).[2] Furthermore, from 1990 to 2019, mortality rates from cardiovascular diseases, which require high ambulance transport rates, increased by 108% in Indonesia and 134% in Malaysia.[3] The growing demand for emergency care, coupled with the aging of populations in both countries, highlights the need for improvement in the systems of emergency care. Studies have shown the current inadequacy of emergency care in Indonesia and Malaysia. A study in Indonesia found that a high proportion of patients with myocardial infarction (62.8%) failed to receive timely reperfusion therapy because they arrived at the emergency department (ED) over 12 hours after symptom onset.[4] Another study in Malaysia showed that the average “door-to-needle” time for acute myocardial infarction (time from ED arrival to initiation of thrombolytic therapy) was approximately 105 minutes, exceeding the golden time of less than 30 minutes to improve outcomes.[5] These findings demonstrate the need for further improvement in emergency care in Indonesia and Malaysia.

In addition to system-level issues, emergency care education in Indonesia and Malaysia has not yet been fully established. Emergency medicine was officially recognized as a specialty in Indonesia in 2017, but the number of certified emergency physicians who have completed the specialty training is still limited. As a result, most emergency care is provided by general practitioners who have not received specialized training.[6] In Malaysia, the number of certified emergency physicians is 7.4 per million population, which is fewer than half of that in Singapore.[1] Additionally, in Malaysia, there are currently no standardized educational courses for prehospital care providers.[7] To make matters worse, several barriers hinder the development of a standardized system of emergency care in these countries, including the lack of a national consensus on ambulance triage protocols, insufficient numbers of ambulances and paramedics, low priority given to ambulances in traffic congestion, prolonged ambulance response times, and low utilization and awareness of ambulance services.[8,9]

In this context, improvements in the emergency care system is a necessity in both Indonesia and Malaysia to reduce preventable deaths and improve public health. Nevertheless, few studies have evaluated what issues should be prioritized within the current system. Our study aims to clarify the current challenges in the emergency care systems in Indonesia and Malaysia as recognized by frontline staff, including physicians, nurses, and medical officers working in the ED.

## Materials & Methods

### Design

This study is an online survey conducted from August to November 2022 using Google Forms (Alphabet Inc. California, USA). A total of five hospitals from Indonesia and six hospitals from Malaysia participated (11 hospitals total). The names and locations of the participating hospitals are shown in **Supplemental Figure 1**. Ethical approval for this study was waived by the central ethics committee of TXP Medical. Co. Ltd, but each participating site obtained ethical approval based on policies of the local ethics committees.

The URL of the online questionnaire was distributed to the ED staff at each participating hospital, including doctors, nurses, and medical workers through the chief investigator at each site. During the study period, staff were not permitted to discuss the content of the survey within their department or communicate with ED staff at other sites about the survey. The purpose of the survey and the respondents’ rights were described on the first page of the questionnaires (See **Appendix**). Only those who agreed with the survey policy and provided informed consent on the first page of the survey were allowed to participate in the survey. Participation was anonymous and voluntary, and no financial incentives were offered for completing the survey.

### Survey settings

Five hospitals from Indonesia and six hospitals from Malaysia participated in the study. The information on the participating hospitals is provided in **Supplemental Table 1**. In Indonesia, the participating hospitals had a median of 778 employees, with four of the five hospitals being secondary hospitals and one being a tertiary hospital. Out of the five hospitals, three were national hospitals and two were private hospitals. The median number of hospital beds was 125 and of ED beds was 25, and a median of 1825 patients was accepted per month. In Malaysia, six tertiary national hospitals participated in the study. Out of the six hospitals, five were national hospitals and one was a private hospital. They had a median of 3750 employees, 1054 beds and 51 ED beds, and received a median of 6130 patients per month.

### Contents of the online survey and measurements

We collected the following information: 1) respondent characteristics (five questions), 2) factors associated with the quality of prehospital care (six questions), 3) factors associated with the quality of emergency care in the ED (eight questions), 4) missing clinical information from prehospital care or ambulance team to the ED (11 questions), and 5) factors associated with the length of patient stay at the ED (six questions). The original questionnaire consisted of 47 questions and was estimated to take approximately 10 minutes to complete (see **Appendix**). Duplicated responses were excluded from the analysis.

This survey consists of two main sections: 1) Respondent Demographic Characteristics, which collected information about the characteristics of the participant, and 2) Survey on the Emergency Department, which aimed to investigate the factors associated with ED management and identify issues recognized by the participants. The questions were initially produced in English with the help of a native English speaker, then translated into the lingua franca in each country: Bahasa Indonesia for Indonesia and Bahasa Malaysia for Malaysia.

### Statistical analysis

The characteristics of the participating hospitals and responders were reported as medians and interquartile ranges (IQRs) for continuous variables and as numbers and percentages (%) for categorical variables. Factors associated with prehospital and emergency care, which were collected using a 10-point Likert scale, were analyzed as medians with IQRs and means and were shown separately for each country, Indonesia and Malaysia.[10,11] Means were used to identify the top three factors associated with prehospital and emergency care. We first describe the results from Indonesia, followed by the results from Malaysia. Finally, we identify shared factors among the top three factors associated with prehospital and emergency care in the two countries. All data analyses were performed using R version 4.2.1 (R Foundation for Statistical Computing, Vienna, Austria).

## Results

## 1. Indonesia

### 1.1 Characteristics of respondents in Indonesia

In Indonesia, we obtained responses from 83 participants from five hospitals. As shown in **Table 1**, 27% were male, mostly aged between 20–29 (59%) followed by 30–39 (29%). Nurses accounted for 58% of respondents, followed by emergency physicians certified as specialists in emergency medicine (22%). Approximately half of the respondents had 1–4 years of experience in the current position (54%). The highest academic qualification among the respondents was an undergraduate degree (51%), followed by a diploma (31%).

**Table 1.** Characteristics of respondents

|  | **Indonesia** | **Malaysia** |
| --- | --- | --- |
| **Characteristic** | **(n=83)** | **(n=109)** |
| Male | 22 (27) | 56 (51) |
| Age (year) |  |  |
| 20-29 | 49 (59) | 38 (35) |
| 30-39 | 24 (29) | 61 (56) |
| 40-49 | 10 (12) | 7 (6) |
| 50-59 | 0 (0) | 3 (3) |
| Occupation |  |  |
| Assistant medical officer | 0 (0) | 41 (38) |
| Emergency physician (certified as a specialist in<br>emergency medicine) | 18 (22) | 2 (2) |
| Emergency physician (non-certified) | 10 (12) | 0 (0) |
| Medical officer | 1 (1) | 47 (43) |
| Nurse | 48 (58) | 19 (17) |
| Pharmacist or health worker in pharmacy | 4 (5) | 0 (0) |
| Other | 2 (2) | 0 (0) |
| Experience in the current position (year) |  |  |
| <1 | 2 (2) | 6 (6) |
| 1-4 | 45 (54) | 36 (33) |
| 5-8 | 13 (16) | 32 (29) |
| >8 | 23 (28) | 35 (32) |
| Highest academic qualification |  |  |
| Diploma | 26 (31) | 57 (53) |
| Master's degree | 2 (2) | 5 (5) |
| Undergraduate degree | 42 (51) | 47 (43) |
| Others | 13 (16) | 0 (0) |

Data in the table are presented as number (percentage)

### 1.2 Factors associated with the quality of prehospital care in Indonesia

All items had a median score of 6 (of 10) or higher (higher score indicates more problematic issue) (**Figure 1a**), indicating that the participants felt that there were several issues with the quality of prehospital care. The most important issue was “inadequate amount of clinical information from the ambulance (**Table 2**)” followed by “inefficient systems to receive and transcribe the information from the ambulance to the hospital medical record” and “inaccurate information from the ambulance.”

**Figure 1.**
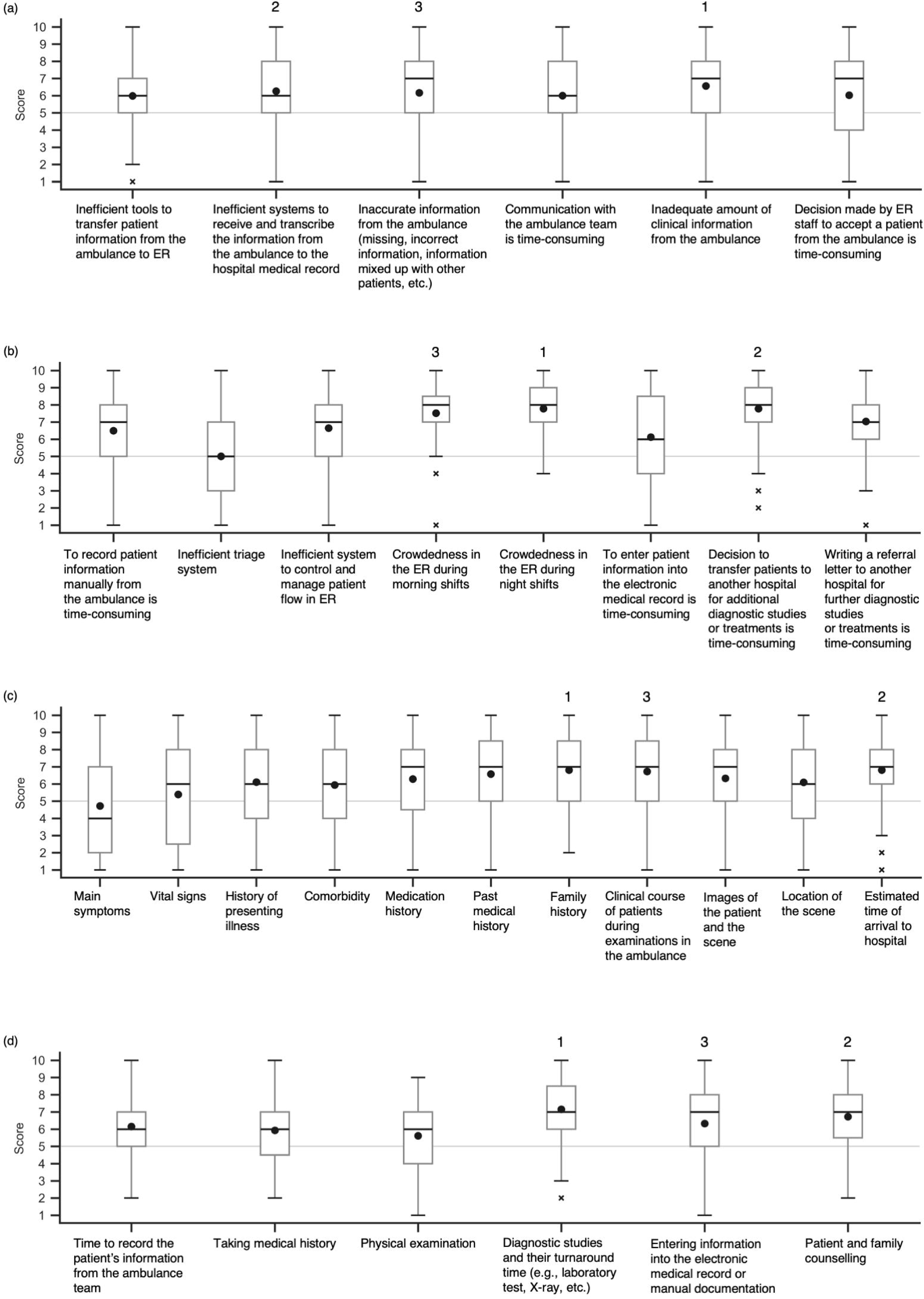
Box plots of survey results for emergency care issues in Indonesia. The lines inside each boxplot represent the median value and the circles inside each boxplot represent the mean value. The top three highest mean values in each boxplot are labeled 1, 2, and 3 above the corresponding boxes. (a) Factors associated with the quality of prehospital care. Scale: 1 = Strongly Disagree, 10 = Strongly Agree. (b) Factors associated with the quality of emergency care in the ED. Scale: for the first question, 1 = Strongly Disagree, 10 = Strongly Agree; for the second question, 1 = Least crowded, 10 = Most crowded; for the last two questions, 1 = Very easy, 10 = Time-consuming. (c) Missing clinical information from prehospital care to the ED. Scale: 1 = Strongly Disagree, 10 = Strongly Agree. (d) Factors associated with the length of patient stay at the ED. Scale: 1 = Very short, 10 = Very long. ED, emergency department

**Table 2.** Top 3 factors associated with the delivery of emergency care

|  | **Indonesia** | **Malaysia** |
| --- | --- | --- |
| **Factors associated with the quality of prehospital care** |  |  |
| Top 1 | Inadequate amount of clinical information from the ambulance (median, 7 [IQR, 5-8]) | Inadequate amount of clinical information from the ambulance (median, 5 [IQR, 4-7]) |
| Top 2 | Inefficient systems to receive and transcribe the information from the ambulance to the hospital medical record (median, 6 [IQR, 5-8]) | Inefficient tools to transfer patient information from the ambulance to ER (median, 5 [IQR, 4-6]) |
| Top 3 | Inaccurate information from the ambulance (missing, incorrect information, information mixed up with other patients, etc.) (median, 7 [IQR, 5-8]) | Inefficient systems to receive and transcribe the information from the ambulance to the hospital medical record (median, 5 [IQR, 4-7]) |
| **Factors associated with the quality of emergency care in the ED** |  |  |
| Top 1 | Crowdedness in the ER during night shifts (median, 8 [IQR, 7-9]) | Crowdedness in the ER during night shifts (median, 8 [IQR, 6-9]) |
| Top 2 | Decision to transfer patients to another hospital for additional diagnostic studies or treatments is time-consuming (median, 8 [IQR, 7-9]) | Decision to transfer patients to another hospital for additional diagnostic studies or treatments is time-consuming (median, 8 [IQR, 6-9]) |
| Top 3 | Crowdedness in the ER during morning shifts (median, 8 [IQR, 7-9]) | Writing a referral letter to another hospital for further diagnostic studies or treatments is time-consuming (median, 7 [IQR, 5-8]) |
| **Missing clinical information from prehospital care to the ED** |  |  |
| Top 1 | Family history (median, 7 [IQR, 5-9]) | Family history (median, 7 [IQR, 5-9]) |
| Top 2 | Estimated time of arrival to hospital (median, 7 [IQR, 6-8]) | Medication history (median, 7 [IQR, 4-8]) |
| Top 3 | Clinical course of patients during examinations in the ambulance (median, 7 [IQR, 5-9]) | Images of the patient and the scene (median, 6 [IQR, 3-9]) |
| **Factors associated with the length of patient stay at the ED** |  |  |
| Top 1 | Diagnostic studies and their turnaround time (e.g., laboratory test, X-ray, etc.) (median, 7 [IQR, 6-9]) | Diagnostic studies and their turnaround time (e.g., laboratory test, X-ray, etc.) (median, 8 [IQR, 7-9]) |
| Top 2 | Patient and family counselling (median, 7 [IQR, 6-8]) | Patient and family counselling (median, 6 [IQR, 5-7]) |
| Top 3 | Entering information into the electronic medical record or manual documentation (median, 7 [IQR, 5-8]) | Entering information into the electronic medical record or manual documentation (median, 5 [IQR, 4-7]) |

Abbreviations: IQR, interquartile range; ER, emergency room; ED, emergency department

### 1.3 Factors associated with the quality of emergency care in the ED in Indonesia

All items had a median score of 6 (of 10) or higher (**Figure 1b**), indicating that the participants felt that there were several issues with the quality of patient care in the ED. The most important issues were “crowdedness in the emergency room (ER) during night shifts” and “decision to transfer patients to another hospital for additional diagnostic studies or treatments is time-consuming” (both with same scores; **Table 2**). In contrast, about half of the participants believed the current triage system was sufficient (a median score of 5 for “inefficient triage system”).

### 1.4 Failure to transfer clinical information from prehospital care to the ED in Indonesia

Most clinical information from the prehospital care or the ambulance team to the ED (e.g., family history, estimated time of arrival to hospital, clinical progress of patient during the ambulance examinations) was had a median score of 6 or higher, which indicates that relevant information was not adequately transferred to the ED, with the exception of information on main symptoms, which was relatively well transferred (median score of 4; **Figure 1c** and **Table 2**). The clinical information with the highest score (least likely to be transferred from prehospital care to the ED) was family history, followed by estimated time of arrival to hospital and clinical course of patients during examinations in the ambulance.

### 1.5 Factors associated with the length of patient stay at the ED in Indonesia

All factors had a score of 6 or higher (**Figure 1d**). Among them, “diagnostic studies and their turnaround time (e.g., laboratory test, X-ray, etc.),” “patient and family counseling,” and “entering information into the electronic medical record or manual documentation” had the highest scores in this order (**Table 2**). These findings suggest that the participants had concerns about various aspects of the ED processes that were causing extended patient stays.

## 2. Malaysia

### 2.1 Characteristics of respondents in Malaysia

In Malaysia, 109 respondents from six hospitals participated (**Table 1**). A majority of the respondents were male (51%), were between 30–39 (56%), and were Medical Officer or Assistant Medical Officer (82%). The respondents’ experience in their current position was equally distributed into 1–4, 5–8, and more than 8 years. The highest academic qualification among the respondents in Malaysia was a diploma (53%), followed by an undergraduate degree (43%).

### 2.2 Factors associated with the quality of prehospital care in Malaysia

All items had a median score of 5 (**Figure 2a**). The most important factor was “inadequate amount of clinical information from the ambulance (**Table 2**)” followed by “inefficient tools to transfer patient information from the ambulance to ER” and “inefficient systems to receive and transcribe the information from the ambulance to the hospital medical record.” Overall, the participants had concerns about various aspects of the quality of patient care in the prehospital setting.

**Figure 2.**
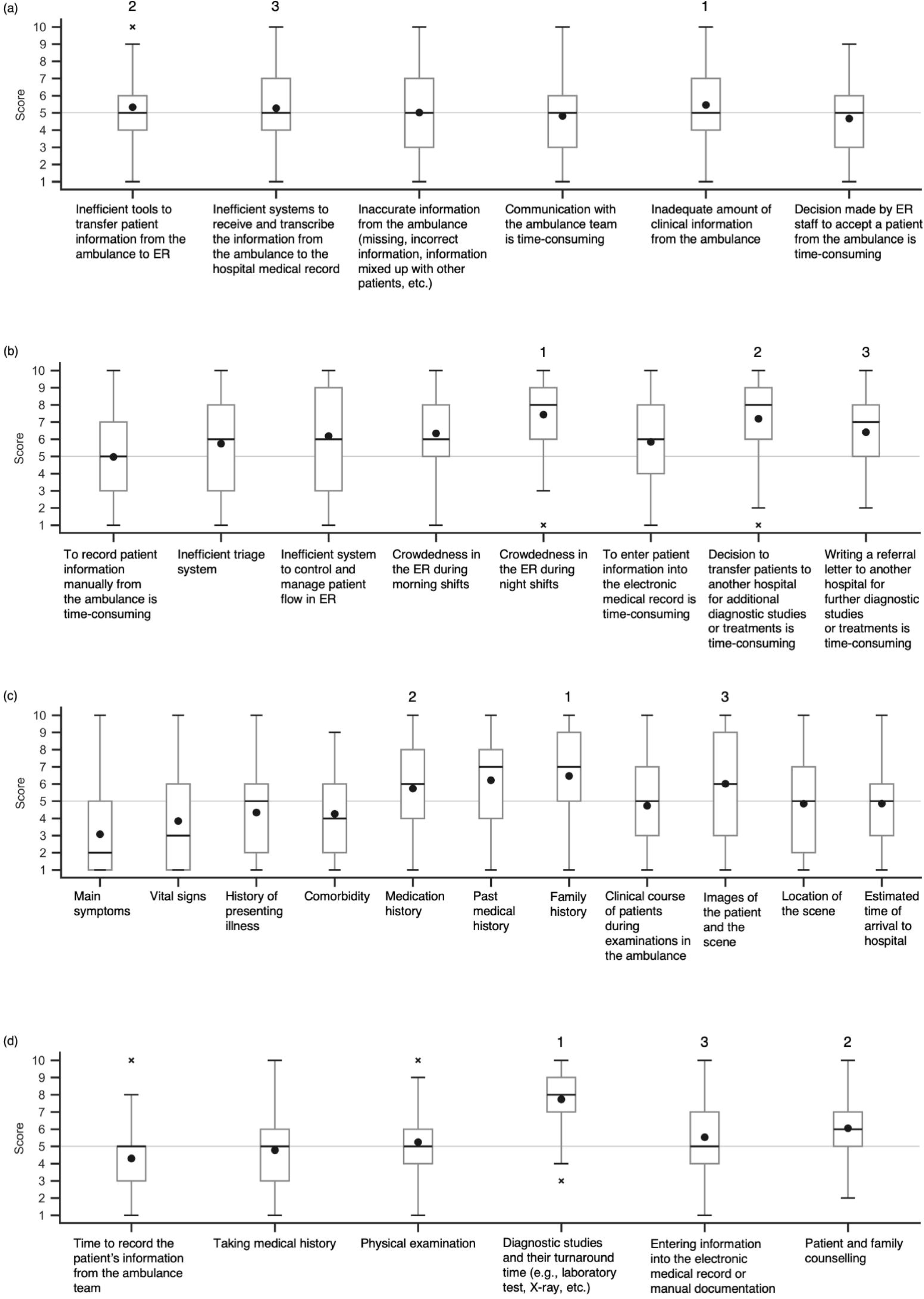
Box plots of survey results for emergency care issues in Malaysia. The lines inside each boxplot represent the median value and the circles inside each boxplot represent the mean value. The top three highest mean values in each boxplot are labeled 1, 2, and 3 above the corresponding boxes. (a) Factors associated with the quality of prehospital care. Scale: 1 = Strongly Disagree, 10 = Strongly Agree. (b) Factors associated with the quality of emergency care in the ED. Scale: for the first question, 1 = Strongly Disagree, 10 = Strongly Agree; for the second question, 1 = Least crowded, 10 = Most crowded; for the last two questions, 1 = Very easy, 10 = Time-consuming. (c) Missing clinical information from prehospital care to the ED. Scale: 1 = Strongly Disagree, 10 = Strongly Agree. (d) Factors associated with the length of patient stay at the ED. Scale: 1 = Very short, 10 = Very long. ED, emergency department

### 2.3 Factors associated with the quality of emergency care in the ED in Malaysia

Most items had a median score of 6 of 10 or higher, reflecting a general perception of these factors as problematic issues (**Figure 2b**). The most important issues were “crowdedness in the ER during night shifts (**Table 2**)” and “decision to refer patients to another hospital for additional diagnostic studies or treatments is time-consuming.” In contrast, about half of the participants felt that the current triage system was sufficient, as indicated by a median score of 5 for “to record patient information manually from the ambulance is time-consuming.”

### 2.4 Failure to transfer clinical information from prehospital care to the ED in Malaysia

Most clinical information from the prehospital care or the ambulance team to the ED (e.g., family history, estimated time of arrival to hospital, clinical course of patients during examinations in the ambulance) had a median score of 6 or higher, which indicates that relevant information was not adequately transferred to the ED, with the exception of information on main symptoms, which was relatively well transferred (**Figure 2c** and **Table 2**). The clinical information with the highest score (least likely to be transferred from prehospital care to the ED) was family history, followed by medical history and images of the patient and the scene.

### 2.5 Factors associated with the length of patient stay at the ED in Malaysia

All factors had a median score of 5 or higher (**Figure 2d**); participants thought there were several factors contributing to prolonged stay in the ED. The highest factors were “diagnostic studies and their turnaround time,” “patient and family counselling,” and “entering information into the electronic medical record or manual documentation” (**Table 2**).

## 3. Factors commonly recognized in both Indonesia and Malaysia

Among the top three factors of each section (factors associated with the quality of prehospital care, factors associated with the quality of emergency care in the ED, missing clinical information from prehospital care to the ED, and factors associated with the length of patient stay at the ED), factors with the highest scores were the same between Indonesia and Malaysia: “inadequate amount of clinical information from the ambulance,” “crowdedness in the ER during night shifts,” “family history,” and “diagnostic studies and their turnaround time,” respectively.

## Discussions

We identified features of current prehospital and emergency care in Indonesia and Malaysia that should be targeted for improvement in the quality of care and the overall healthcare system. This is the first survey to uncover the issues or factors affecting the quality of emergency care as recognized by frontline staff in EDs in Indonesia and Malaysia. The survey results showed that in both Indonesia and Malaysia, important factors affecting the quality of patient care in the prehospital setting were “inadequate amount of clinical information from the ambulance” and “inefficient systems to receive and transcribe the information from the ambulance to the hospital medical record.” Ambulance teams are tasked with a multitude of responsibilities, including patient care and examination, gathering patient information, communicating with the hospital ED, and monitoring the patient during transport.[12] The challenge is to balance the collection of clinical information with the time available for patient care.[13] In severe cases, the priority is to provide immediate care and treatment, which may limit the time available for additional information collection.[13]

To address these issues, a novel and innovative approach for data collection and transfer could be a solution. For instance, using smartphone applications to transfer data from ambulance teams to hospitals could reduce communication time with the ED.[14] Additionally, the use of optical character recognition or voice recording to capture clinical information (e.g., vital signs) from ambulance teams or the automatic transfer of data from the vital sign monitors in the ambulance to the ED could reduce the risk of missing information. The crowdedness in the ER during night shifts was rated as the highest issue affecting quality of patient care in the ED. Although optical character recognition and voice recording could streamline the process of inputting information into electrical medical records,[14] it may not necessarily improve the turnaround time for diagnostic studies or the time required for patient and family counseling. Given that “diagnostic studies and their turnaround time (e.g., laboratory test, X-ray, etc.)” was recognized as a factor associated with the length of patient stay at the ED, more robust approaches that enable immediate data sharing across departments in the hospital might be beneficial in alleviating crowding.

Interestingly, this study demonstrated that issues prioritized by staff were consistent between the two countries. The best approach may be to first explore the solution that can address these issues. In addition, these factors may be prevalent in other developing countries, so solutions may be effective in these countries as well. In contrast, some factors varied across the two countries, suggesting that factors affecting the quality of prehospital and emergency care could vary widely across countries. This also highlights the importance of tailoring interventions based on the specific background and situation of each setting and that a one-size-fits-all solution is unlikely to be effective.[15,16]

This study should be interpreted in light of several limitations. First, our survey was based on the perceptions of staff working in the hospital ED, and there may be significant discrepancies between the identified factors and in actual clinical practice. As a result, relationships between the factors identified in this survey and patient outcomes should not be inferred from this study alone. However, we are planning to conduct additional studies to further investigate this, as patient outcomes are of utmost importance. Despite this limitation, we were able to capture valuable perspectives from frontline staff that had not been previously obtained and we hope that these findings can inform future discussions and interventions aimed at improving patient care. Second, the number of participating hospitals and respondents was limited, and extrapolation of the results to clinical practice or policymaking should be done with caution. However, conducting large-scale surveys through questionnaires can be quite challenging in developing countries where large-scale survey panels are not yet in place. Despite these difficulties, this study attempted to overcome these challenges and can still detect potential priority areas for intervention.

## Conclusions

This survey identified the challenges faced by frontline staff in providing quality emergency care in Indonesia and Malaysia. Our results identified top priority factors to be addressed in prehospital and emergency care, such as lack of information from ambulance services, overcrowding in the ER, and prolonged time of input into medical records. Based on the findings, implementing targeted interventions may help promote better health outcomes, taking into account the specific context of each country.

## Author contribution statement

Akio Tokita: contributed to draft manuscript and literature search, and performed formal analysis.

Hanako Nunokawa: contributed to draft manuscript and literature search.

Keibun Liu: conceptualized the study, collected and analyzed data, interpreted data, and provided supervision.

Yuta Iwamoto: conceptualized the study, acquired funding, and provided supervision. Tomohiro Sonoo: acquired funding and provided supervision.

Konan Hara: performed formal analysis and provided supervision. Mikio Nakajima: contributed to methodology and provided supervision.

Kiyomitsu Fukaguchi: contributed to methodology and provided supervision. Takanori Takeda: acquired and curated data.

Amirudin bin Sanip, Dafsah A. Juzar, Gurjeet Singh a/l Harvendhar Singh, Lukito Condro, Monalisa, Muhammad Abdus-Syakur Bin Abu Hasan, Nik Hisamuddin NA Rahman, Orizanov Mahisa, Ramdinal Aviesena Zairinal, and Mohd Khairulizwan Ramli: acquired, curated, and validated data.

Tadahiro Goto: conceptualized the study and provided supervision.

Mohd Idzwan bin Zakaria: conceptualized the study, curated data, and provided supervision.

## Contributors

Corona Rintawan, Rachmat Ardiyanzah PG, and Imanda Dyah Rahmadani from Muhammadiyah Hospital Lamongan, Lamongan, Indonesia

Rinni Nuraini from Emergency Department, Hermina Depok Hospital, Depok, Indonesia Bobi Prabowo from RSUD Kanjuruhan Kabupaten Malang, Kepanjen, Indonesia Bambang Widyantoro from Department of Cardiology and Vascular Medicine, Faculty of Medicine Universitas Indonesia - National Cardiovascular Center Harapan Kita, Jakarta, Indonesia

Mohammad Aizuddin Azizah Ariffin, Mohd Hafyzuddin Md Yusuf, and Aidawati Bustam from Academic Unit Trauma and Emergency, Faculty of Medicine, University Malaya, Kuala Lumpur, Malaysia

Sarah Zhahir, Faizal Amri, Tan Toh Leong, En Abdul Karim Mustafa, Pn Rafidah Md Kamal, Mohammad Nur Kamal Effendy Husein, and Nurul Saadah Ahmad from Department of Emergency Medicine, Universiti Kebangsaan Malaysia, Kuala Lumpur, Malaysia

Mohd Faiz Mohd Shukri and Mohd Shaharudin Shah Che Hamzah from Hospital Universiti Sains Malaysia, Health Campus, Kubang Kerian, Kota Bharu Kelantan, Malaysia Mohamed Alwi Bin Haji Abdul Rahman and Nabil Muhammad Al Kuddoos from Hospital Selayang, Batu Caves, Malaysia

Zainalabidin bin Mohamed from Hospital Tengku Ampun Afzan, Kuantan, Malaysia S. M. Wazien Wafa B. S. Saadun Tarek Wafa and Mohd Nadzrul Shah Bin Mohd Junit from Kulliyyah of Medicine, International Islamic University Malaysia, Kuantan, Malaysia

## Data availability statements

The datasets generated and/or analyzed during the current study are available from the corresponding author on reasonable request.

