## Supplemental material for "The current quality and requirements of prehospital and emergency care in Indonesia and Malaysia: a cross-sectional multicenter survey"

**Supplementary material**

**Supplemental Table 1. Characteristics of participating hospitals**

| **Characteristic** | **Indonesia**  **(n=5)** | **Malaysia**  **(n=6)** |
| --- | --- | --- |
| Total number of employees in the hospital | 778 [700, 1039] | 3750 [3550, 4025] |
| Level of healthcare |  |  |
| Secondary | 4 (80) | 0 (0) |
| Tertiary | 1 (20) | 6 (100) |
| Type of hospital |  |  |
| National hospital | 3 (60) | 5 (83) |
| Private hospital | 2 (40) | 1 (17) |
| Number of hospital beds | 125 [113, 138] | 1054 [969, 1127] |
| Number of beds in ER | 25 [21, 29] | 51 [34, 56] |
| Number of patients per month transferred to ER | 1825 [1336, 1922] | 6130 [6020, 6930] |
| Number of ER patients per month transferred by local ambulance from the scene | 163 [82, 420] | 934 [778, 1138] |
| Number of ER patients per month transferred by private car from the scene | 800 [530, 1150] | 5000 [5000, 5500] |
| Number of ER patients per month transferred from other medical facilities (patient transport between hospitals) | 152 [84, 270] | 125 [39, 300] |

Data in the table are presented as median [interquartile range] or number (percentage)

Abbreviations: ER, emergency room

**Supplemental Figure 1. Participating hospitals**

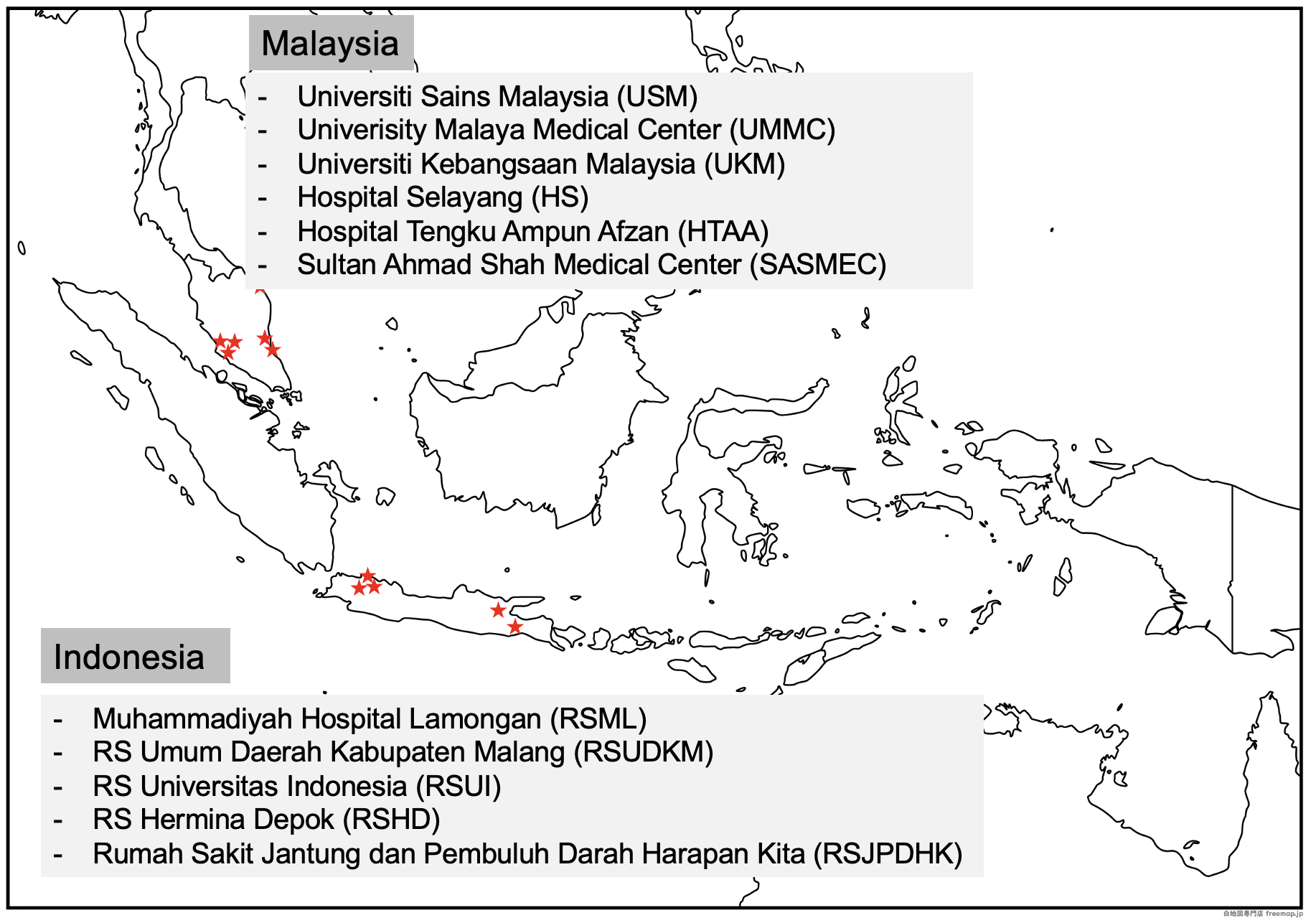

Pre-intervention phase

**Online Survey on the current status and standard of emergency medicine and trauma services at the emergency department in Indonesia**

To all who are interested in participating in the pilot project with TXP Medical Co. Ltd. and Department of Emergency Medicine in Indonesia.

The objective of this research is to investigate the current status of the Indonesian emergency medicine and trauma care system before the implementation of the NSER mobile in prehospital care. Your contribution to this study is invaluable for the improvement of the emergency department (ED).

Please read through the description of the survey below and click "Participate" at the bottom of the page to proceed to the survey.

**Preliminary Confirmation on study participation**

1. This survey consists of two main sections, "Respondents Demographic Characteristics" and "Survey on the emergency department," with a total of 20 questions (estimated time: 10 minutes).
2. There will be no patient information involved in this study only your honest opinions on the current ED. We appreciate your candid responses.
3. Participation in this survey is voluntary. This survey can be dropped at any time if you decide not to participate.
4. Survey respondents are guaranteed the right to refuse to respond and will not be disadvantaged or compelled to respond if they change their minds and wish to terminate their response midway through the survey.
5. Ethical approval (number) was obtained on “date”.

If you have any questions or concerns about this survey or its ethics, please do not hesitate to contact the principal investigator. We will sincerely try to answer and resolve your questions.

 Principal Investigator of this survey:

Keibun Liu (TXP Medical Co. Ltd):

Is this the first time you answer this questionnaire? (This question aims to prevent duplicate responses from the same person.)

 □Yes □No (Choose "Yes" to continue, "No" to end)

For those who selected “Yes”, if you agree to participate after reading the above, please click **“Agree to participate”** at the bottom of this web page to start the questionnaire. If you do not agree to participate in this survey, please click **“Disagree to participate”** at the bottom to close this web page.

 □Agree to participate □Disagree to participate

**[Respondents demographic characteristics]**

Please write the first 5 characters of your email address you received this survey from. This information will be used as a unique key for future surveys. If you have less than 5 characters in your email address, please write as many as possible.

For example, if your email address is “keibun@”, the answer would be “keibu”.

If you email address is “keib@”, the answer would be “keib”.

1. Age (years)

- ~19 years old
- 20~29 years old
- 30~39 years old
- 40~49 years old
- 50~59 years old
- 60 years old or older

2. Gender

- Male
- Female

3. Occupation

- Emergency physician (Certified)
- Emergency physician (Non-certified)
- Medical officer
- Assistant Medical Officer (AMO)
- Nurse
- Emergency Medical Technician (EMT)
- Other (free text)

4. Years of experience in your current position (years)

5. Current position:
a. AMO U29 or equivalent
b. Nurse U29 or equivalent
c. AMO U32 and above
d. Nurse U32 and above
e. Medical officer any grades
f. Emergency physician

6. Highest academic qualification
a. Diploma
b. Undergraduate degree
c. Master’s degree
d. PhD

**［Survey on the emergency department (ED)]**

1. **Outpatient** care in the ED provided by your center is in accordance to national standard.

Non-critical cases are managed in ED according to national standard

| 1 | 2 | 3 | 4 | 5 | 6 | 7 | 8 | 9 | 10 |
| --- | --- | --- | --- | --- | --- | --- | --- | --- | --- |

Strongly disagree	Strongly Agree

Critical cases are managed in ED according to national standard

| 1 | 2 | 3 | 4 | 5 | 6 | 7 | 8 | 9 | 10 |
| --- | --- | --- | --- | --- | --- | --- | --- | --- | --- |

Strongly disagree	Strongly Agree

2. What are the issues related to the ED system?

(PHC: pre-hospital care)

- Decision made by call center staff to accept a patient from the pre-hospital care (PHC) is time-consuming

| 1 | 2 | 3 | 4 | 5 | 6 | 7 | 8 | 9 | 10 |
| --- | --- | --- | --- | --- | --- | --- | --- | --- | --- |

Strongly Disagree	Strongly Agree

- Inadequate clinical information from the pre-hospital care (PHC)

| 1 | 2 | 3 | 4 | 5 | 6 | 7 | 8 | 9 | 10 |
| --- | --- | --- | --- | --- | --- | --- | --- | --- | --- |

Strongly Disagree	Strongly Agree

- To record patient information manually from the pre-hospital care (PHC) is time-consuming

| 1 | 2 | 3 | 4 | 5 | 6 | 7 | 8 | 9 | 10 |
| --- | --- | --- | --- | --- | --- | --- | --- | --- | --- |

Strongly Disagree	Strongly Agree

Preparing medications and equipment to treat medical emergencies prior to patient arrival is common (e.g., thrombolysis for stroke protocol, streptokinase for acute myocardial infarction

| 1 | 2 | 3 | 4 | 5 | 6 | 7 | 8 | 9 | 10 |
| --- | --- | --- | --- | --- | --- | --- | --- | --- | --- |

Strongly Disagree	Strongly Agree

- Inefficient system to control and manage patient flow in ED

| 1 | 2 | 3 | 4 | 5 | 6 | 7 | 8 | 9 | 10 |
| --- | --- | --- | --- | --- | --- | --- | --- | --- | --- |

Strongly Disagree	Strongly Agree

- ED overcrowding/ access block is common on a daily basis

| 1 | 2 | 3 | 4 | 5 | 6 | 7 | 8 | 9 | 10 |
| --- | --- | --- | --- | --- | --- | --- | --- | --- | --- |

Strongly Disagree	Strongly Agree

- Inefficient triage system

| 1 | 2 | 3 | 4 | 5 | 6 | 7 | 8 | 9 | 10 |
| --- | --- | --- | --- | --- | --- | --- | --- | --- | --- |

Strongly Disagree	Strongly Agree

- To enter patient information into the electronic medical record is time-consuming

| 1 | 2 | 3 | 4 | 5 | 6 | 7 | 8 | 9 | 10 |
| --- | --- | --- | --- | --- | --- | --- | --- | --- | --- |

Strongly Disagree	Strongly Agree

- Others (free text)

3. The quality of patient information received from the pre-hospital care (PHC) to the ED is adequate/enough.

| 1 | 2 | 3 | 4 | 5 | 6 | 7 | 8 | 9 | 10 |
| --- | --- | --- | --- | --- | --- | --- | --- | --- | --- |

Strongly Disagree	Strongly Agree

4. What information are commonly missing in pre-hospital care (PHC) report? (Click all that apply)

- Main symptoms

| 1 | 2 | 3 | 4 | 5 | 6 | 7 | 8 | 9 | 10 |
| --- | --- | --- | --- | --- | --- | --- | --- | --- | --- |

Strongly Disagree	Strongly Agree

- Vital signs

| 1 | 2 | 3 | 4 | 5 | 6 | 7 | 8 | 9 | 10 |
| --- | --- | --- | --- | --- | --- | --- | --- | --- | --- |

Strongly Disagree	Strongly Agree

- History of presenting illness

| 1 | 2 | 3 | 4 | 5 | 6 | 7 | 8 | 9 | 10 |
| --- | --- | --- | --- | --- | --- | --- | --- | --- | --- |

Strongly Disagree	Strongly Agree

- Comorbidity

| 1 | 2 | 3 | 4 | 5 | 6 | 7 | 8 | 9 | 10 |
| --- | --- | --- | --- | --- | --- | --- | --- | --- | --- |

Strongly Disagree	Strongly Agree

- Past medical history

| 1 | 2 | 3 | 4 | 5 | 6 | 7 | 8 | 9 | 10 |
| --- | --- | --- | --- | --- | --- | --- | --- | --- | --- |

Strongly Disagree	Strongly Agree

- Medication history

| 1 | 2 | 3 | 4 | 5 | 6 | 7 | 8 | 9 | 10 |
| --- | --- | --- | --- | --- | --- | --- | --- | --- | --- |

Strongly Disagree	Strongly Agree

- Family history

| 1 | 2 | 3 | 4 | 5 | 6 | 7 | 8 | 9 | 10 |
| --- | --- | --- | --- | --- | --- | --- | --- | --- | --- |

Strongly Disagree	Strongly Agree

- Clinical Progress of patient during pre-hospital care (PHC)

| 1 | 2 | 3 | 4 | 5 | 6 | 7 | 8 | 9 | 10 |
| --- | --- | --- | --- | --- | --- | --- | --- | --- | --- |

Strongly Disagree	Strongly Agree

- Images of the patient and scene

| 1 | 2 | 3 | 4 | 5 | 6 | 7 | 8 | 9 | 10 |
| --- | --- | --- | --- | --- | --- | --- | --- | --- | --- |

Strongly Disagree	Strongly Agree

- Location of scene

| 1 | 2 | 3 | 4 | 5 | 6 | 7 | 8 | 9 | 10 |
| --- | --- | --- | --- | --- | --- | --- | --- | --- | --- |

Strongly Disagree	Strongly Agree

- Estimated time of arrival to hospital

| 1 | 2 | 3 | 4 | 5 | 6 | 7 | 8 | 9 | 10 |
| --- | --- | --- | --- | --- | --- | --- | --- | --- | --- |

Strongly Disagree	Strongly Agree

Others (free text)

5. From the following options, rate the possible problems in communication from pre-hospital care (PHC) to the ED/ call center.

- Inefficient tools to transfer patient information from pre-hospital care (PHC) to ED PRIOR to arrival (i.e., during transportation)

| 1 | 2 | 3 | 4 | 5 | 6 | 7 | 8 | 9 | 10 |
| --- | --- | --- | --- | --- | --- | --- | --- | --- | --- |

Strongly Disagree	Strongly Agree

- Inefficient systems to receive and transcribe the patient information from pre-hospital care (PHC) to the hospital medical record

| 1 | 2 | 3 | 4 | 5 | 6 | 7 | 8 | 9 | 10 |
| --- | --- | --- | --- | --- | --- | --- | --- | --- | --- |

Strongly Disagree	Strongly Agree

- Inaccurate pre-hospital care (PHC) information is common (missing, incorrect information is sometimes given, information is mixed up with other patients, etc.)

| 1 | 2 | 3 | 4 | 5 | 6 | 7 | 8 | 9 | 10 |
| --- | --- | --- | --- | --- | --- | --- | --- | --- | --- |

Strongly Disagree	Strongly Agree

- Communication with pre-hospital care (PHC) team is time-consuming

| 1 | 2 | 3 | 4 | 5 | 6 | 7 | 8 | 9 | 10 |
| --- | --- | --- | --- | --- | --- | --- | --- | --- | --- |

Strongly Disagree	Strongly Agree

- Others (free text)

6. When do you prepare the medications and equipment necessary for medical emergency prior to patient arrival to the ED? (Click all that apply)

- Immediately after receiving the call from pre-hospital care (PHC)
- After the patient arrives to the ED
- Not prepared
- Others (Free text)

7. The quality of the current patient triage system in the ED is…

| 1 | 2 | 3 | 4 | 5 | 6 | 7 | 8 | 9 | 10 |
| --- | --- | --- | --- | --- | --- | --- | --- | --- | --- |

Not working at all	Excellent

8. How are pre-hospital care (PHC) information handled upon patient arrival to the receiving hospital?

- The information will not be stored in the hospital database
- The information will be stored in the hospital paper based database
- The information will be stored in the electronic medical record
- Others (free text)

9. The current effort to enter patient information into the electronic medical record in the ED is…

| 1 | 2 | 3 | 4 | 5 | 6 | 7 | 8 | 9 | 10 |
| --- | --- | --- | --- | --- | --- | --- | --- | --- | --- |

Very easy	Time-consuming

10. How crowded is the ED during these shift periods?

Morning shift (i.e., 0700–1400hours)

| 1 | 2 | 3 | 4 | 5 | 6 | 7 | 8 | 9 | 10 |
| --- | --- | --- | --- | --- | --- | --- | --- | --- | --- |

Least crowded	Most crowded

Afternoon shift (i.e., 1400–2100hours)

| 1 | 2 | 3 | 4 | 5 | 6 | 7 | 8 | 9 | 10 |
| --- | --- | --- | --- | --- | --- | --- | --- | --- | --- |

Least crowded	Most crowded

Night shift (i.e., 2100-0700hours)

| 1 | 2 | 3 | 4 | 5 | 6 | 7 | 8 | 9 | 10 |
| --- | --- | --- | --- | --- | --- | --- | --- | --- | --- |

Least crowded	Most crowded

11. The decision to decant patient to another hospital for further investigations or treatments from the ED is…

| 1 | 2 | 3 | 4 | 5 | 6 | 7 | 8 | 9 | 10 |
| --- | --- | --- | --- | --- | --- | --- | --- | --- | --- |

Very easy	Time-consuming

12. The current effort involved in writing a referral letter, when the patient needs to be transported to another hospital for further investigations or treatments from the ED is…

| 1 | 2 | 3 | 4 | 5 | 6 | 7 | 8 | 9 | 10 |
| --- | --- | --- | --- | --- | --- | --- | --- | --- | --- |

Very easy	Time-consuming

13. The patient’s length of stay in ED is…

| 1 | 2 | 3 | 4 | 5 | 6 | 7 | 8 | 9 | 10 |
| --- | --- | --- | --- | --- | --- | --- | --- | --- | --- |

Very short	Very long

14. From the following options, rate each factor that may contribute to the patient’s ED length of stay

- Time to record the patient’s information from the pre-hospital care (PHC) team

| 1 | 2 | 3 | 4 | 5 | 6 | 7 | 8 | 9 | 10 |
| --- | --- | --- | --- | --- | --- | --- | --- | --- | --- |

Very short	Very long

- Taking medical history

| 1 | 2 | 3 | 4 | 5 | 6 | 7 | 8 | 9 | 10 |
| --- | --- | --- | --- | --- | --- | --- | --- | --- | --- |

Very short	Very long

- Physical examination

| 1 | 2 | 3 | 4 | 5 | 6 | 7 | 8 | 9 | 10 |
| --- | --- | --- | --- | --- | --- | --- | --- | --- | --- |

Very short	Very long

- Investigations and their turn around time (e.g., laboratory test, X-ray, etc.)

| 1 | 2 | 3 | 4 | 5 | 6 | 7 | 8 | 9 | 10 |
| --- | --- | --- | --- | --- | --- | --- | --- | --- | --- |

Very short	Very long

- Entering information into the electronic medical record or manual documentation

| 1 | 2 | 3 | 4 | 5 | 6 | 7 | 8 | 9 | 10 |
| --- | --- | --- | --- | --- | --- | --- | --- | --- | --- |

Very short	Very long

- Patient and family counselling

| 1 | 2 | 3 | 4 | 5 | 6 | 7 | 8 | 9 | 10 |
| --- | --- | --- | --- | --- | --- | --- | --- | --- | --- |

Very short	Very long

- Other (free text)
